# Medical domain knowledge in domain-agnostic generative AI

**DOI:** 10.1101/2022.01.10.22269025

**Authors:** Jakob Nikolas Kather, Narmin Ghaffari Laleh, Sebastian Foersch, Daniel Truhn

## Abstract

The text-guided diffusion model GLIDE (Guided Language to Image Diffusion for Generation and Editing) is the state of the art in text-to-image generative artificial intelligence (AI). GLIDE has rich representations, but medical applications of this model have not been systematically explored. If GLIDE had useful medical knowledge, it could be used for medical image analysis tasks, a domain in which AI systems are still highly engineered towards a single use-case. Here we show that the publicly available GLIDE model has reasonably strong representations of key topics in cancer research and oncology, in particular the general style of histopathology images and multiple facets of diseases, pathological processes and laboratory assays. However, GLIDE seems to lack useful representations of the style and content of radiology data. Our findings demonstrate that domain-agnostic generative AI models can learn relevant medical concepts without explicit training. Thus, GLIDE and similar models might be useful for medical image processing tasks in the future - particularly with additional domain-specific fine-tuning.

## Generative models: from GANs to GLIDE

Generative artificial intelligence (AI) models can create synthetic images which are very close to or indistinguishable from real images. Broadly used techniques in this field are generative adversarial networks (GANs)^1^ and variational autoencoders (VAEs)^2^, of which many variants exist. These generative models can create synthetic images, in an unconditional way or conditional to a defined set of classes. Both approaches require dedicated training to a specific domain, for example x-ray images. In this domain, conditional generative models can be trained on large datasets with multiple disease categories, enabling them to synthesize images in any of these categories. Recently, generative AI approaches have been extended by coupling language processing models and image processing models.^3^ These multimodal models can be used for image generation from text prompts.^4^ Trained on millions of image-text pairs, such algorithms build rich internal representations of many facets of our world. Thus, these models can be used for zero-shot image generation, i.e. synthesizing images of categories which were not explicitly represented in the training set.^4^ The state of the art in multimodal generative AI is the text-guided diffusion model GLIDE^5^, which recently outperformed another recent generative model, DALL-E^4^. GLIDE can generate realistic and complex images across many domains and has demonstrated the capability of logical reasoning. Potential medical applications of these models have not been systematically explored.

## Generative models in medicine

In medical applications of AI, generative models have been used in multiple data types, including radiology^6^, histopathology^7,8^ and endoscopy^9^. Synthetic data generated by such AI systems is hailed as a promising approach for data augmentation, data sharing and explainability in medical AI.^10^ However, an important limitation of generative AI in medicine is the limited scope of such specialized systems which often need to be laboriously trained to generate images in a single narrow domain.^11^ This problem could be addressed by a domain-agnostic approach such as GLIDE: If training on vast amounts of unselected text-image pairs were sufficient to generate useful synthetic medical images, this could massively improve the adoption of such models in medical applications. However, it is unclear if training a large AI model on unselected text-image pairs scraped from the internet also conveys useful medical knowledge. Currently, the reality in medical AI is that AI systems focus on a narrow niche with a single type of data and be validated thoroughly in this particular niche.^11^ However, such “narrow” AI systems are arguably less useful than generalist models, and it might be impossible to create and validate AI systems for every conceivable niche application in medicine.

## How much medical domain knowledge is encoded in GLIDE?

We experimentally investigated if GLIDE has plausible representations on medical styles and medical concepts with a focus on cancer research and oncology (**Figure 1**). We use the publicly available, somewhat restricted version of GLIDE, which was released in December 2021 and showed comparable performance to the previous state-of-the-art model, DALL-E, which was released in early 2021. A limitation of the public GLIDE model is that photographs of people were removed from the training dataset. However, the general concept of diseases or representations of medical image data is not necessarily negatively affected by such filtering. Using medicine-related text prompts for GLIDE, we found that images of high photographic quality were generated, but observed some striking confusions: For example, the term “Grave’s disease” resulted in a photograph of a gravestone (**Figure 2A**). Also, the text prompt “A histopathological image of the brain” resulted in a macroscopic cross-section of the brain shown in the style of a histopathology image (**Figure 2A**). Furthermore, many images, especially illustrations had illegible text in the image, pointing to limitations in the available model which presumably did not learn robust typesetting during training. In general, the model seemed to have understood some key concepts, but combined them in ways that were sometimes surprising, such as a heart made of blood vessels, an X-ray image of a tree-like structure in the brain (**Figure 1**). Obvious confusions were more prevalent in images generated without CLIP guidance (**Figure 2B-C**), but also present in some images generated with CLIP guidance (**Figure 2D**). This is in contrast to findings made in the original publication of GLIDE by Nichol et al.^5^, which demonstrated that GLIDE without CLIP guidance results in higher quality images. Indeed, we subjectively reproduced Nichol et al.’s finding for non-medical text prompts, in which the subjective photographic quality was higher for GLIDE without CLIP guidance (**Figure 2E**). We conclude that although GLIDE without CLIP guidance is superior for non-medical text prompts, CLIP guidance improves the results. In addition, we compared short text prompts (**Suppl. Figure 1B**) with detailed text prompts (**Suppl. Figure 1B**) and found that detailed prompts generally improved style and content.

**Figure 1:**
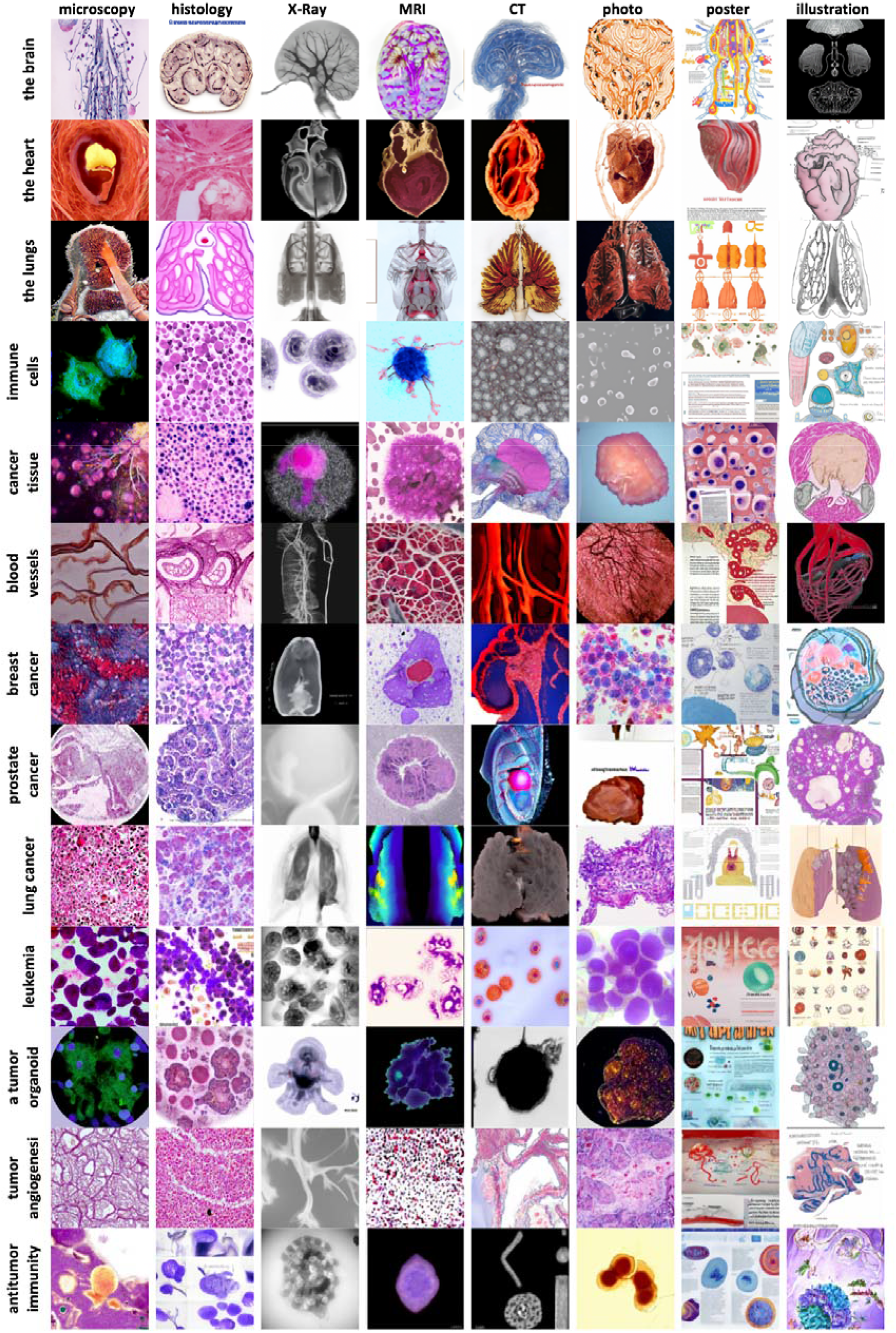
Example images by target category and modality. All images were generated with a CLIP-conditioned GLIDE. One example was chosen from eight instances per category.

**Figure 2:**
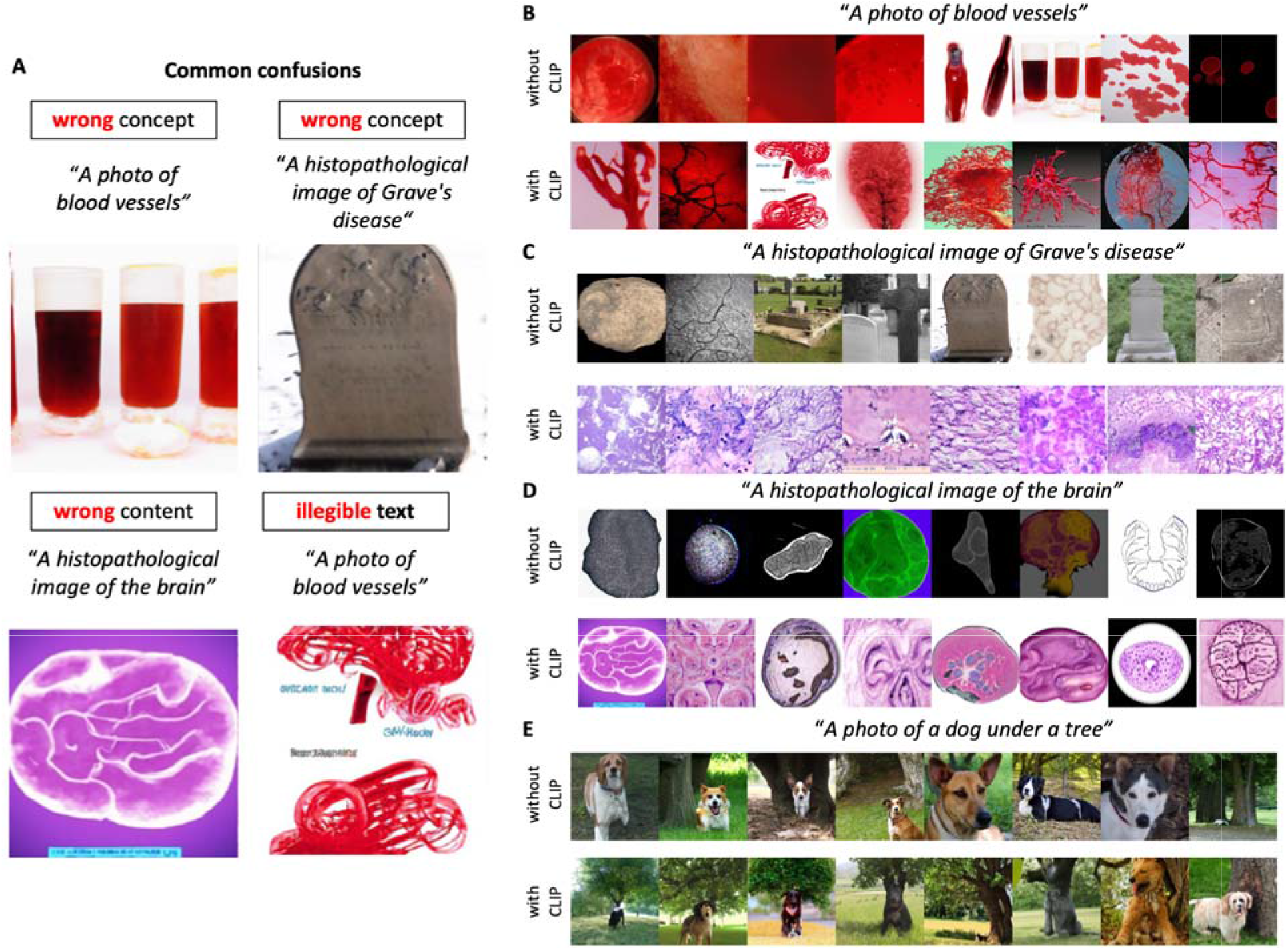
Common confusions of the model. (A) Example images of common confusions observed in our study, (B-E) Generated images without and with CLIP guidance for four text prompts. For each prompt, eight random images are shown and these images are not cherry-picked.

## Systematic evaluation of style and content in GLIDE-generated images

How good is GLIDE in generating a correct style and correct content for medicine-related text prompts? To quantitatively analyze this, we generated hundreds of images with GLIDE (with CLIP guidance) and asked three medical experts to classify the correctness of style and content shown on these images (**Suppl. Table 1**). Style and content were interpreted by a pathologist in training, a radiologist and an internist on an ordinal rating scale from 0 to 4 (0=completely wrong,1=hints of correct, 2=clear signs of correct, 3=mainly correct, 4=perfectly correct) in a blinded way. They were presented with four randomly generated images and rated the best style and content in these images (**Suppl. Figure 2**, data and code in https://github.com/KatherLab/synth-user-study). Specifically, we generated images in multiple styles: microscopy, histology, x-ray, MRI, CT, photograph, poster and illustration. Using this approach, we found that GLIDE reached a high median score of 3 out of 4 for generating the style of histopathology images, scientific posters and scientific illustrations (**Suppl. Figure 3A**). Regarding the content in the synthetic images, GLIDE reached a moderate median score of 2 out of 4 for microscopy, histopathology, photography and illustrations (**Suppl. Figure 3B**). The style and content of radiology images (x-ray, magnetic resonance [MR] and computed tomography [CT] images) was poor and scored significantly (p<0.05) lower than all other categories (**Suppl. Figure 3C-D**). Together, these data demonstrate that GLIDE was able to synthesize realistic scientific posters and illustrations with plausible content, which is not surprising given that these types of data were presumably part of the training dataset. More interesting, however, is the observation that GLIDE was able to generate histopathology images with convincing style and sometimes even with partly accurate content. These results demonstrate that domain-agnostic generative AI models such as GLIDE could potentially be used for generation of histopathology images for research purposes. For radiology applications, however, GLIDE falls markedly short of the quality of specialized generative AI models^6^, but could potentially be used after domain-specific fine-tuning. In general, text-to-image generative models such as GLIDE could be used in medical image analysis applications, solving the problem of researchers having to train generative models on every single niche domain.

## Challenges and outlook

GLIDE and CLIP are very large models. Training them from scratch requires much more computing power and data than most researchers have access to. Our experiments show that the publicly available models already have useful medical information in their latent representations. Thus, researchers could use these models without modifications or fine-tune on specific tasks. A potential option for broad re-training on a large set of tasks in a medical field (e.g. histopathology) would be to couple GLIDE/CLIP with automated data mining strategies as recently suggested by Schaumberg et al.^12^ However, re-training on specific tasks negates the benefit of having broadly applicable models with zero-shot classification and generation capabilities. Future studies should investigate the tradeoff between domain specificity and zero-shot performance. In general, such models could be used for a plethora of tasks, including education and training, data anonymization, data augmentation and discovery of new morphological associations and potentially of biological mechanisms. This could be achieved by having sensible outputs for prompts like “A histology image of a patient who benefits from immunotherapy” or “An MRI image of a patient who should be treated with a statin” and then by gradually escalating the text prompts towards more difficult tasks. An important area for future research will be optimal prompt engineering, i.e. identifying text (or image) prompts for optimal synthetic images for the desired application. These prompts might differ by use case. For example, for education and training purposes, an exaggeration of characteristic details might be desirable. On the other hand, for some data anonymization and augmentation purposes, it might be wise to avoid characteristic features in the model output. Furthermore, a broader validation of the generated data by various domain experts is crucial. While classical generative AI architectures are confined to the specific categories which were present in the training dataset, the zero-shot capabilities of GLIDE and similar models could make it possible to use massive unlabeled datasets for medical knowledge generation. Potentially, medical-domain-specific CLIP guidance could further improve image content substantially. All this could conceivably expand the applicability and usefulness of generative AI in medicine in the future.

## Data Availability

All data produced are available online at https://github.com/KatherLab/synth-user-study

## Additional information

### Author contributions

All authors conceived the idea, analyzed the data and wrote the manuscript. JNK performed the data analysis and created the visualizations.

