## Supplementary Material for "Medical domain knowledge in domain-agnostic generative AI"

**supplementary data**

### Supplementary Figures

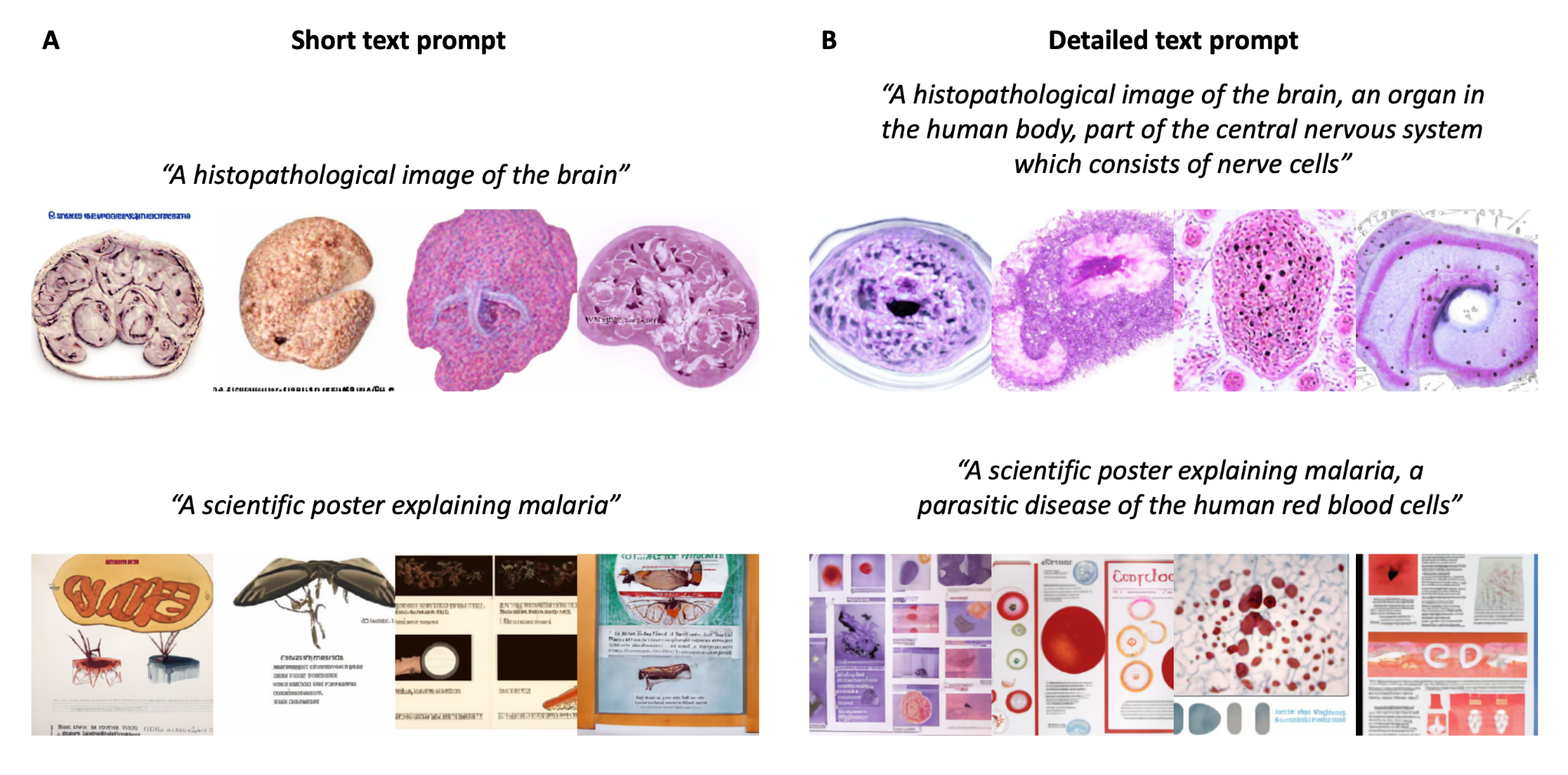

**Suppl. Figure 1: Text prompt engineering for GLIDE with CLIP guidance.** (A) simple text prompts,
(B) detailed text prompts. Four random images are shown per category, images are not cherry-picked.

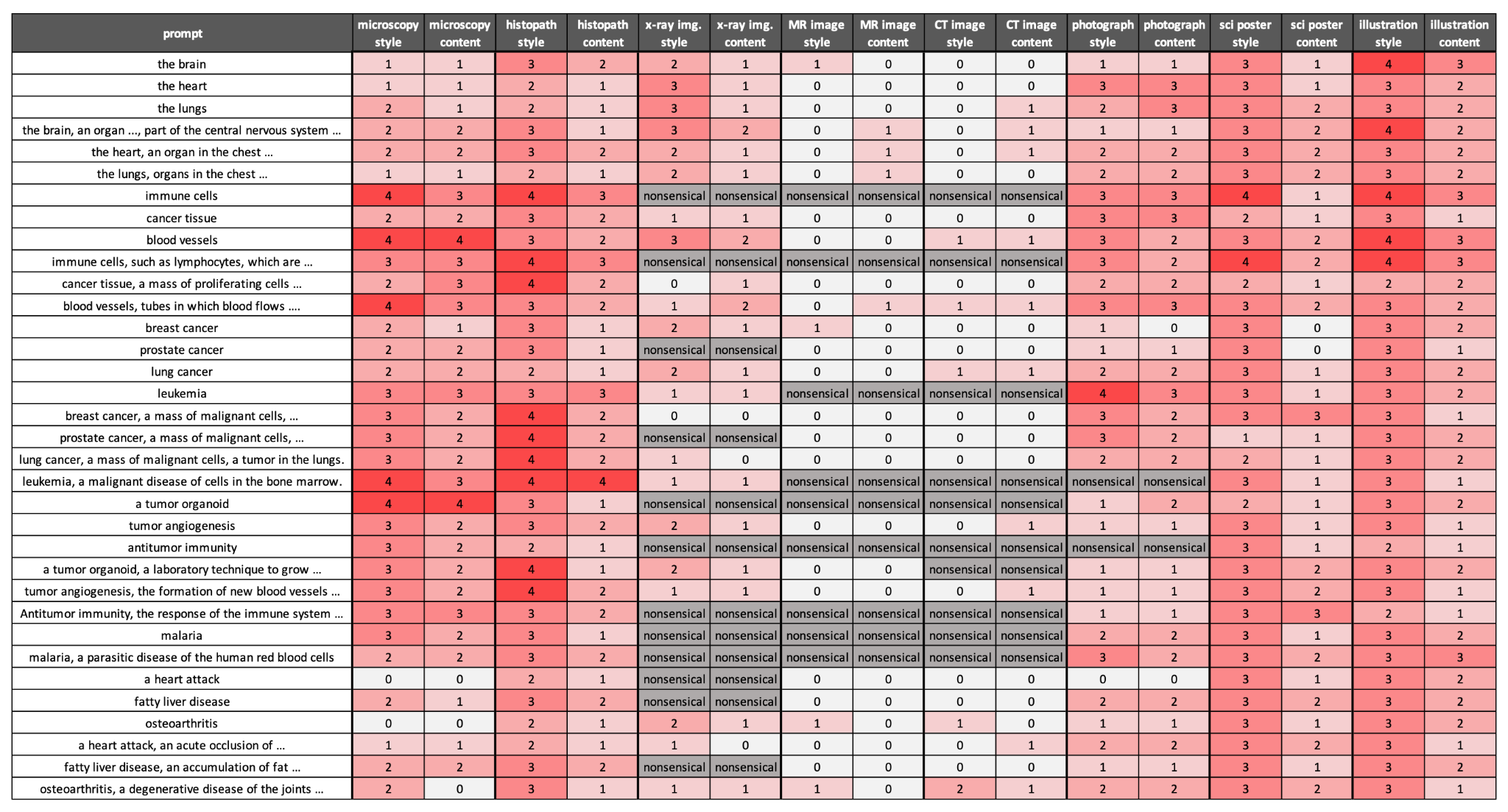

**Suppl. Figure 2: Results of observer study.** Three observers scored style and content on a numerical rating scale ranging from 0 to 4.
All values are reported as median, “nonsensical” if one observer scored the category as “nonsensical”.

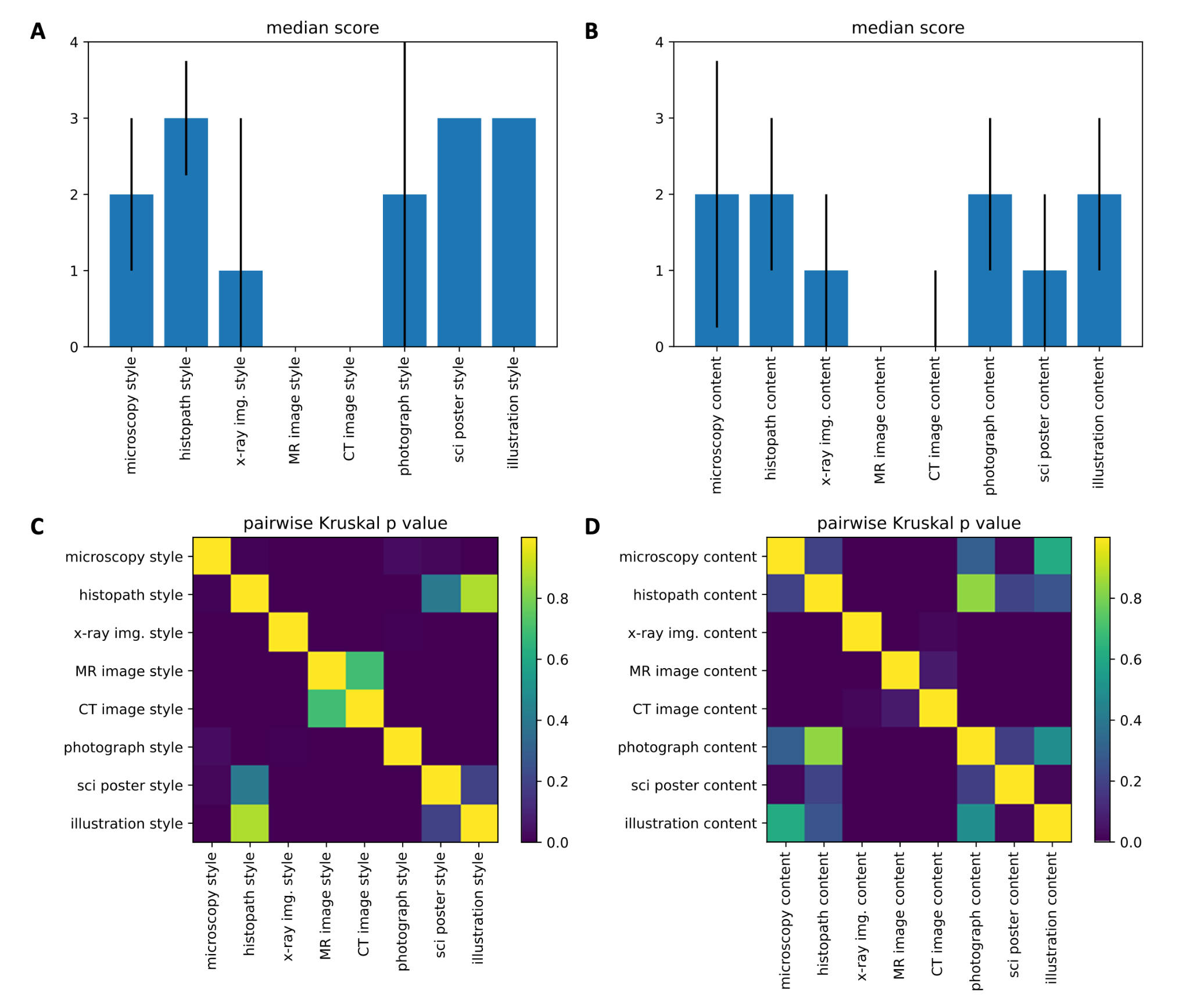

**Suppl. Figure 3: Summary of observer study.** Median +/- interquartile range (25th to 75th percentile),
**(A)** Median scores (all observers, all items) in each category for style and **(B)** for content. **(C)** Pairwise Kruskal p value for
comparison of median scores for all items between categories, for style and **(D)** for content. Categories which were scored as
“nonsensical” by any observer were set to 0 (lowest score).

### Supplementary Tables

| Part 1 of prompts | 1. A histopathological image of 2. A microscopic image of 3. An x-ray image of 4. A magnetic resonance image of 5. A computed tomography image of 6. A schematic drawing of 7. A photo of | |
| --- | --- | --- |
| Part 2 of prompts  … organs | 1. the brain 2. the heart 3. the lungs | 1. the brain, an organ in the human body, part of the central nervous system which consists of nerve cells. 2. the heart, an organ in the chest of the human body which consists of muscle cells. 3. the lungs, organs in the chest of the human body which consists of airways and alveoli. |
| …  cancer tissues | 1. immune cells 2. cancer tissue 3. blood vessels | 1. immune cells, such as lymphocytes, which are part of the human immune system. 2. cancer tissue, a mass of proliferating cells which invade the surrounding tissue. 3. blood vessels, tubes in which blood flows in the human body. |
| …  cancer diseases | 1. breast cancer 2. prostate cancer 3. lung cancer 4. leukemia | 1. breast cancer, a mass of malignant cells, a tumor in the breast. 2. prostate cancer, a mass of malignant cells, a tumor in the prostate. 3. lung cancer, a mass of malignant cells, a tumor in the lungs. 4. leukemia, a malignant disease of cells in the bone marrow. |
| …  cancer research | 1. a tumor organoid 2. tumor angiogenesis 3. antitumor immunity | 1. a tumor organoid, a laboratory technique to grow multicellular spheres of tumor cells. 2. tumor angiogenesis, the formation of new blood vessels which nurture tumor cells in a cancer. 3. Antitumor immunity, the response of the immune system against cancer cells. |
| … non-tumor diseases | 1. malaria 2. a heart attack 3. fatty liver disease 4. osteoarthritis | 1. a heart attack, an acute occlusion of the coronary arteries in the heart. 2. fatty liver disease, an accumulation of fat in the cells of the liver in the body. 3. osteoarthritis, a degenerative disease of the joints in the human body. |

**Suppl. Table 1: All text prompts used to generate the images.** Left: short text prompts, Right: detailed text prompts.
